# First Data-Set on SARS-CoV-2 Detection for Istanbul Wastewaters in Turkey

**DOI:** 10.1101/2020.05.03.20089417

**Authors:** Bilge Alpaslan Kocamemi, Halil Kurt, Sabri Hacıoglu, Cevdet Yaralı, Ahmet Mete Saatci, Bekir Pakdemirli

**Author notes:** **Bilge Alpaslan Kocamemi:** Methodology, Data Curation, Formal analysis, Writing - Original Draft, Preparation, Writing - Review & Editing, Visualization, Supervision, Project administration. **Halil Kurt:** Validation, Verification, Data Curation, Writing - Review & Editing. **Sabri Hacıoğlu:** Investigation, **Cevdet Yaralı:** Validation, **Ahmet Mete Saatçı:** Conceptualization, Writing - Review & Editing, Supervision, Project administration. **Bekir Pakdemirli**: Resources, Funding acquisition.

## Abstract

Severe acute respiratory syndrome coronavirus 2 (SARS-CoV-2) started in Wuhan, China, in December 2019 and became a global pandemic [1]. By 26 April 2020, more than 2.9 million people were infected by SARS-CoV-2 and over 203 thousand people lost their life globally. By 26 April 2020, 107773 confirmed cases were reported in Turkey with 2706 deaths. Majority of the cases in Turkey has been observed in Istanbul. In the world, the duration of availability of SARS-CoV-2 was found to be significantly longer in stool samples than in respiratory and serum samples [2]. SARS-CoV-2 was detected in wastewaters in Australia [3], Netherlands [4], USA [5], France [6], Spain [7] and USA [8] by using different virus concentration techniques. In this work, Istanbul metropole with 65 % of Covid-19 cases was chosen as the pilot city. On the 21st of April 2020, 24-hr composite samples were collected from the Ambarli, Pasakoy and Kadikoy wastewater treatment plants (WWTP). On the 25^th^ of April 2020, more wastewater samples were taken from Terkos, Buyukcekmece, Baltalimani and Tuzla WWTPs. These wastewater treatment plants were selected among 81 plants in Istanbul in order to take representative samples from 4 different districts of Istanbul according to the severity of Covid-19 cases, like very serious, serious, moderate and mild. Grab samples were also collected from Bagcilar and Kartal manholes located nearby the pandemic hospitals on April 21^st^, 2020. Polyethylene glycol 8000 (PEG 8000) adsorption [5] SARS-Cov-2 concentration method was used for SARS-CoV-2 concentration after optimization. Real time RT-PCR diagnostic panel validated by US was used to quantify SARS-CoV-2 RNA in raw sewage taken from the inlets of treatment plants and manholes. Five samples out of seven from wastewater and all samples from manholes were tested positive. SARS-CoV-2 in raw sewage from Ambarli, Pasakoy, Kadikoy, Terkos, Buyukcekmece, Baltalimani and Tuzla WWTPs were found as 8.26×10^3^, 1.80×10^4^, ND, ND, 3.73×10^3^, 4.95×10^3^, 2.89×10^3^, respectively. The Bagcilar and Kartal manholes nearby pandemic hospitals exhibited 4.49×10^4^ and 9.33×10^4^, respectively. SARS-CoV-2 virus titers of manhole were higher than those of inlet of WWTPs. The observed copy numbers were presented against the number of Covid-19 cases coming to the WWTP per treatment plant capacity. Quantitative measurements of SARS-CoV-2 in wastewater can be used as a tool in wastewater-based epidemiology (WBE) and it can provide information about SARS-CoV-2 distribution in wastewater of various districts of Istanbul which exhibit different scores of Covid-19 cases. The distribution of epidemy was followed not only with blood test but with wastewater monitoring. This may allow us to identify the districts not exhibiting many Covid-19 cases, but under high risk. Continuous monitoring of wastewater for SARS-Cov-2 may provide an early warning signs before an epidemy starts in case of infection resurge.

## Value of the Data

- The dataset reports the SARS-CoV-2 detection results in wastewater of Istanbul with 15.5 million inhabitants and a very high population density (2987 persons/km2). Hence, the dataset provides information about SARS-CoV-2 distribution in wastewater at various districts having serious, moderate and mild cases and presents Covid-19 case numbers against wastewater SARS-CoV-2 titers.
- As being the seventh systematic study in the world, the dataset presented is quite beneficial for the governors trying to fight with Covid-19 around the world. They will have chance to adapt the methodology to their own countries and monitor the epidemic not only with the blood test but also wastewater monitoring. They may have chance to catch the districts not exhibiting too many cases but under risk.
- This surveillance will indicate a correlation the Covid-19 cases with SARS-CoV-2 RT-qPCR results in wastewater. It has quite potential for verifying the reported number of Covid-19 cases with the real situation. Additionally, continuous monitoring of wastewater for SARS-CoV-2 may provide an early warning sign before an epidemy starts in case of infection resurge.

## Data Description

RT-qPCR raw data and standard curve graphs produced by Biorad CFX96 thermal cycle (Biorad Laborateries, Montreal, Quebec), copy number quantification summary, CQ (CT) results and standard curve results exported from Biorad CFX Manager Version 3.1, CQ were submitted with the Supplementary Data.

SARS-CoV-2 copy numbers per liter and calculated indexes in wastewater samples were summarized in Table 1 and shown in Figure 1 together with case number observed at the sampling district against influent flowrate of WWTPs. The slope of the standards SARS-CoV-2 spike protein gene assay was −3.251 and -with an amplification efficiency of 103%. SARS-CoV-2 virus genome was detected in wastewater samples. Five samples out of seven from wastewater and all samples from manholes were tested positive. SARS-CoV-2 titers were ranged 10^2^ to 10^4^ in WWTPs influent samples. SARS-CoV-2 in raw sewage from Ambarli, Pasakoy, Kadikoy, Terkos, Buyukcekmece, Baltalimani and Tuzla WWTPs were found as 8.26×10^3^, 1.80×10^4^, ND, ND, 3.73×10^3^, 4.95×10^3^, 2.89×10^3^, respectively. The Bagcilar and Kartal manholes nearby pandemic hospitals exhibited 4.49×10^4^ and 9.33×10^4^, respectively. SARS-CoV-2 virus titers of manhole were higher than inlet of WWTPs. Terkos wastewater sample has the highest Case number (person)/WWTP flow (m3/d), but SARS-CoV-2 virus was not detected.

**Table 1.** SARS-CoV-2 RT-qPCR results for Istanbul wastewaters

| Sampling Point | Cq value | Cq SD | RT-qPCR results | RT-qPCR SD | Virus titer per liter | Virus titer per liter SD | slope | Reaction efficiency % | R<sup>2</sup> |
| --- | --- | --- | --- | --- | --- | --- | --- | --- | --- |
| Terkos WWTP | ND | ND | ND | ND | ND | ND | -3.25 | 103 | 0.99 |
| Ambarli WWTP | 38.37 | 1.38 | 6.89E+02 | 5.83E+02 | 8.26E+03 | 7.00E+03 | -3.25 | 103 | 0.99 |
| Pasakoy WWTP | 37.23 | 1.11 | 1.50E+03 | 1.03E+03 | 1.80E+04 | 1.24E+04 | -3.25 | 103 | 0.99 |
| Kadikoy WWTP | ND | ND | ND | ND | ND | ND | -3.25 | 103 | 0.99 |
| Baltalimani WWTP | 38.818 | 0.44 | 4.12E+02 | 1.28E+02 | 4.95E+03 | 1.53E+03 | -3.25 | 103 | 0.99 |
| Buyukcekmece WWTP | 39.183 | 0.1 | 3.11E+02 | 2.23E+01 | 3.73E+03 | 2.67E+02 | -3.25 | 103 | 0.99 |
| Tuzla WWTP | 39.54 | NA | 2.41E+02 | NA | 2.89E+03 | NA | -3.25 | 103 | 0.99 |
| Bagcilar Manhole | 35.91 | 0.98 | 3.75E+03 | 2.83E+03 | 4.49E+04 | 3.40E+04 | -3.25 | 103 | 0.99 |
| Kartal Manhole | 34.667 | 0.36 | 7.78E+03 | 1.95E+03 | 9.33E+04 | 2.34E+04 | -3.25 | 103 | 0.99 |

ND: not detected, NA: not applicable

**Figure 1.**
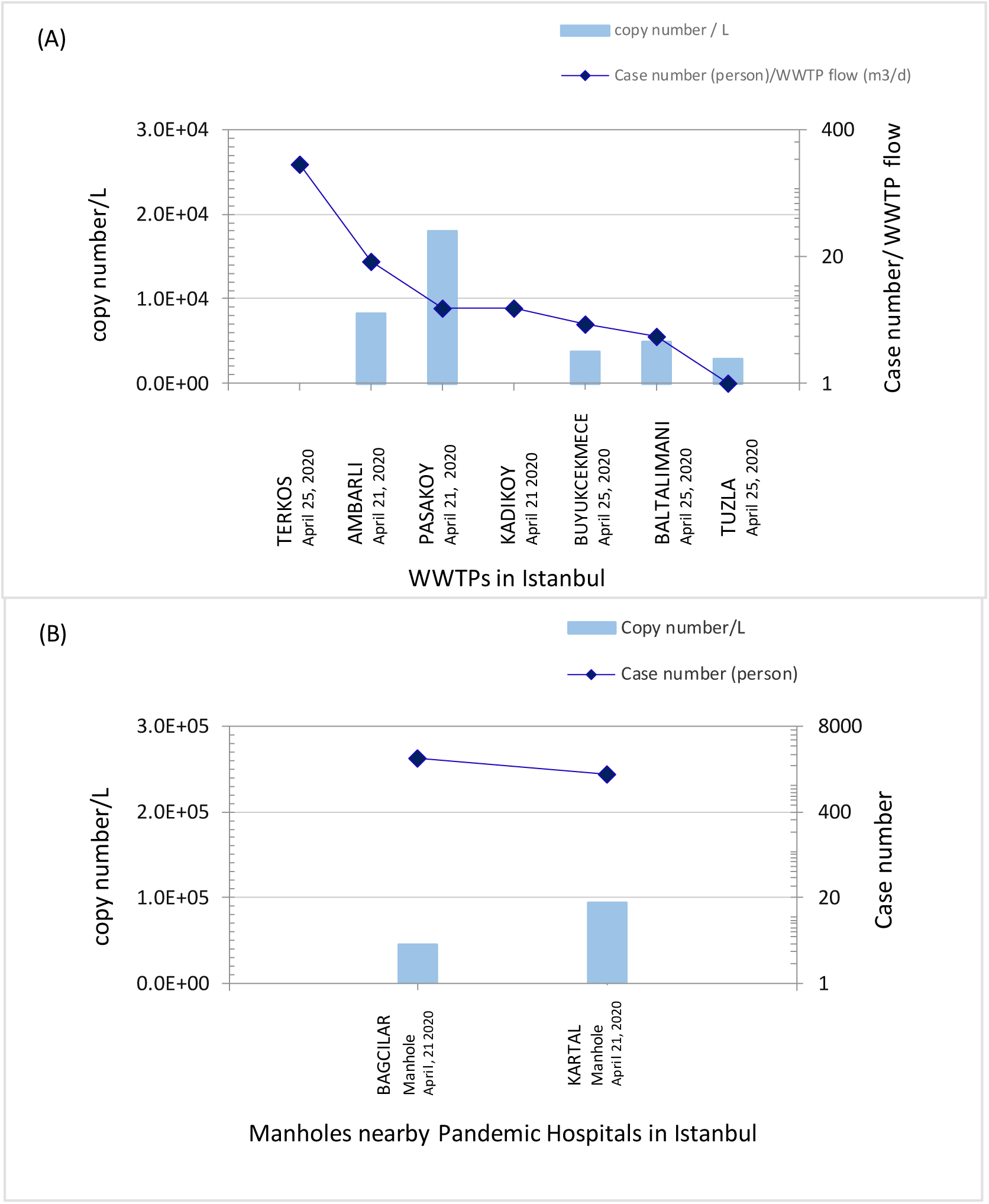
SAR-Cov-2 Levels in Istanbul wastewaters (a) at the inlet of WWTPs, (b) at the manholes nearby pandemic hospitals

## Experimental Design, Materials and Methods

### 2.1 Sampling

The wastewater samples were collected from inlets of seven WWTPs in Istanbul, Turkey and 2 manholes nearby Istanbul epidemic hospitals in 250 ml sterilized bottles. Figure 2 presents a map indicating the first confirmed SARS-CoV-2 detections in the world together with the detection points in Istanbul WWTPs and manholes in Turkey. The map demonstrates sampling points together with the reported Covid-19 cases in the districts of the WWTPs and manholes. The description of sampling points is summarized in Table 2.

**Figure 2.**
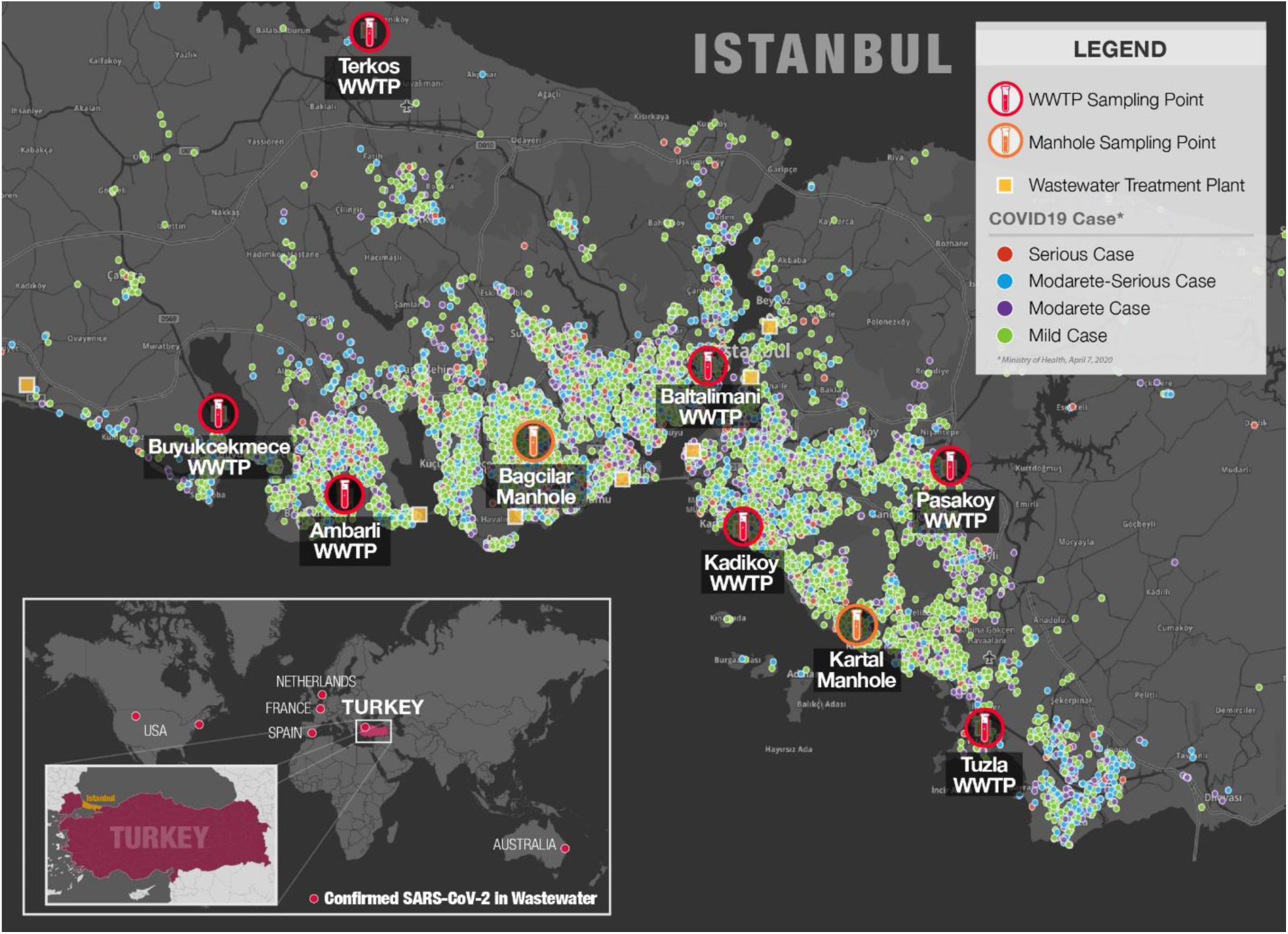
SARS-CoV-2 detection studies in wastewater around the world and in Turkey (Cases from https://geomatic.org/koronovirus on 21st April, 2020.)

**Table 2.** SARS-CoV-2 wastewater detection points in Istanbul WWTPs

| Sampling Point | Coordinate | Type of WWTP | Current Flowrate m3/d | Covid-19 Cases around the district (on 8<sup>th</sup> Ap, 2020) | Index Covid-19 Case / Flowrate |
| --- | --- | --- | --- | --- | --- |
| Terkos WWTP | 41.303137, 28.674010<br>(41°18'11.3"N, 28°40'26.4"E) | Activated Sludge | 1792 | 320 | 179 |
| Ambarli WWTP | 40.988571, 28.683843<br>(40°59'39.0"N, 28°40'58.1"E) | Advanced Nutrient removal | 342715 | 6045 | 18 |
| Pasakoy WWTP | 41.015882, 29.273205<br>(41°00'53.7"N, 29°16'21.8"E) | Advanced Nutrient removal | 167690 | 943 | 6 |
| Kadikoy WWTP | 40.988305, 29.019468<br>(40°59'18.0"N, 29°01'12.5"E) | Preliminary Treatment | 397408 | 2354 | 6 |
| Baltalimani WWTP | 41.096601, 29.050261<br>(41°05'47.8"N, 29°03'00.9"E) | Preliminary Treatment | 183522 | 927 | 5 |
| Buyukcekmece WWTP | 41.046486, 28.553018<br>(41°02'47.4"N, 28°33'10.9"E) | Advanced Nutrient removal | 66277 | 284 | 4 |
| Tuzla WWTP | 40.825605, 29.295600<br>(40°49'32.2"N, 29°17'44.2"E) | Advanced Nutrient removal | 357841 | 1042 | 3 |
| Bagcilar Manhole | 41.029145, 28.870616<br>(41°01'47.9"N, 28°52'14.8"E) | N/A | N/A | 2658 | N/A |
| Kartal Manhole | 40.917165, 29.175142<br>(40°54'58.3"N, 29°10'31.4"E) | N/A | N/A | 1481 | N/A |

### 2.2 SARS-CoV-2 concentration

Efficient and reliable virus concentration and recovery from wastewater are the most important step in SARS-CoV-2 detection. So far, ultracentrifugation [6], Polyethylene glycol 8000 (PEG 8000) adsorption [5], electronegative membrane [3] and ultrafiltration [3, 4, 8] methods were used for SARS-CoV-2 concentration from wastewater samples. The work was first started with the use of Amicon^®^ Ultra-15 with 10KDa cutoff ultrafiltration units (Millipore Corporation, Billerica, MA, USA) to concentrate virus [4]. 250 ml wastewater samples were collected from the inlet of Ankara Tatlar WWTP (39.903239, 32.459330, 39°54’11.7”N, 32°27’33.6”E) which collects wastewater from central Ankara. Large particles such as Bacteria and debris were removed by centrifugation at 3200G for 45 mins (Eppendorf 5810R plus swing-out rotor# A4-62; Eppendorf AG, Hamburg, Germany) without brake. Then, a volume of 250 ml supernatant was filtered using ultrafiltration units by centrifugation at 3200G for 25-40 minutes. (Eppendorf 5810R plus swing-out rotor# A4-62; Eppendorf AG, Hamburg, Germany). The concentrate (200 – 600 μl) was collected from the filter unit. Total RNA was extracted with The QIAamp cador Pathogen Mini Kit (Qiagen, Hilden, Germany) following the manufacturer’s protocols. The QuantiNova Pathogen +IC Kit (Qiagen, Hilden, Germany) were added to samples to evaluate PCR inhibition following the manufacturer’s protocols. However, a high degree of inhibition was observed in the samples. Expected Ct value of internal control was shifted from 30 to 35. As an alternative to UF method, the polyethylene glycol (PEG) adsorption method [10] was tried. Briefly, 250 ml wastewater samples were collected from the inlet of Ambarli WWTP in Istanbul. Large particles such as Bacteria and debris were removed by centrifugation at 3000G for 45 mins (Eppendorf 5810R plus swing-out rotor# A4-62; Eppendorf AG, Hamburg, Germany) without brake. The supernatant was sequentially filtered with 0.45 μm and 0.2 μm filters to remove remaining particles. 5X PEG Stock Solution were prepared by dissolving 100 g of PEG 8000 (Nordic Chemical Solutions, Norway) and 17.5 g of NaCl in 200 ml distilled water. The pH was adjusted to pH 7.0-7.2. Final stock solution had 50 % (w/v) PEG 8000 and 1.5 M NaCl. The solution was sterilized with 0.2 μm filter. Filtrate was mixed thoroughly with PEG 8000 (10% w/v) by shaking for 1 minute. The mixture was incubated at 4°C at 60 rpm for overnight. Following to incubation, virus was precipitated with Centrifuge (Eppendorf 5810R plus swing-out rotor#F34-6-38; Eppendorf AG, Hamburg, Germany) at 4°C at 5700 G for 120 minutes. Supernatant was removed carefully without disturbing the pellets. Pellets was re-suspended with 600 μl RNA free water and stored at −80°C for a long time. A volume of 200 μl virus concentrate was used for total RNA extraction as described above. Due to high community risk of using SARS-CoV-2 virus, 300 μl of 10^5^ copy/ml surrogate avian coronavirus of Infectious Bronchitis Virus were added our samples in order to evaluate the virus recovery efficiency of PEG 8000 adsorption method. Based on RT-QPCR results, 1-1.5 log virus titer loss were observed after PEG 8000 adsorption and RNA isolation.

### 2.3. Quantitative reverse transcription PCR (RT-qPCR)

Primers taqman probe sets targeting SARS-CoV-2 RdRp gene were used in this study [9]. Serial dilution of SARS-CoV-2 RdRp gene were used as a standard to quantify results. RT-qPCR analysis was performed in 20 μL QuantiNova Pathogen +IC Kit (Qiagen, Hilden, Germany) contained 0.8 nM of forward primer and reverse primer, 0.25 nM probe and 5 μL of template RNA. The RT-qPCR assays were performed using a Bio-Rad CFX96 thermal cycler (Bio-Rad Laboratories, Montreal, Quebec).

## Ethics Statement

The work did not involve any human subject and animal experiments.

## Data Availability

The authors confirm that the data supporting the findings of this study are available within the article and its supplementary materials.

## Acknowledgments

This work was financed by Republic of Turkey, Ministry of Agriculture and Forestry.

The authors wish to acknowledge the Turkish Water Institute (SUEN) for the coordination and execution of this study. We wish to express our appreciation to the Ministry’s Veterinary Control Centeral Research Institute for their hard work and rapid analysis of the wastewater samples. We thank to DSI (State Hydraulic Works) for their logistic support for intercity sample transportation. We also thank to ISKI (Istanbul Water and Sewerage Administration) for their cooperation and efforts to collect and preserve the sewage samples rapidly. Our special thanks to Dr. Esra Erdim from Marmara University Department of Environmental Engineering for her contribution to gather and collate up to date information about the worldwide studies on SARS-CoV-2 in wastewater.

## References

[1] F. Wu, S. Zhao, B. Yu, Y.M. Chen, W. Wang, Z.G. Song, Y. Hu, Z.W. Tao, J.H. Tian, Y.Y. Pei, M.L. Yuan, Y.L. Zhang, F.H. Dai, Y. Liu, Q.M. Wang, J.J. Zheng, L. Xu, E.C. Holmes, Y.Z. Zhang, A new coronavirus associated with human respiratory disease in China, Nature. (2020). https://doi.org/10.1038/s41586-020-2008-3.

[2] Y. Wu, C. Guo, L. Tang, Z. Hong, J. Zhou, X. Dong, H. Yin, Q. Xiao, Y. Tang, X. Qu, L. Kuang, X. Fang, N. Mishra, J. Lu, H. Shan, G. Jiang, X. Huang, Prolonged presence of SARS-CoV-2 viral RNA in faecal samples, Lancet Gastroenterol. Hepatol. (2020). https://doi.org/10.1016/S2468-1253(20)30083-2.

[3] W. Ahmed, N. Angel, J. Edson, K. Bibby, A. Bivins, J.W. O’Brien, P.M. Choi, M. Kitajima, S.L. Simpson, J. Li, B. Tscharke, R. Verhagen, W.J.M. Smith, J. Zaugg, L. Dierens, P. Hugenholtz, K. V. Thomas, J.F. Mueller, First confirmed detection of SARS-CoV-2 in untreated wastewater in Australia: A proof of concept for the wastewater surveillance of COVID-19 in the community, Sci. Total Environ. (2020) 138764. https://doi.org/10.1016/J.SCITOTENV.2020.138764.

[4] G. Medema, L. Heijnen, G. Elsinga, R. Italiaander, A. Brouwer, Presence of SARS-Coronavirus-2 in sewage, MedRxiv. (2020) 2020.03.29.20045880. https://doi.org/10.1101/2020.03.29.20045880.

[5] F. Wu, A. Xiao, J. Zhang, X. Gu, W.L. Lee, K. Kauffman, W. Hanage, M. Matus, N. Ghaeli, N. Endo, C. Duvallet, K. Moniz, T. Erickson, P. Chai, J. Thompson, E. Alm, SARS-CoV-2 titers in wastewater are higher than expected from clinically confirmed cases, MedRxiv. (2020) 2020.04.05.20051540. https://doi.org/10.1101/2020.04.05.20051540.

[6] S. Wurtzer, V. Marechal, J.-M. Mouchel, L. Moulin, Time course quantitative detection of SARS-CoV-2 in Parisian wastewaters correlates with COVID-19 confirmed cases, MedRxiv. (2020) 2020.04.12.20062679. https://doi.org/10.1101/2020.04.12.20062679.

[7] W. Randazzo, P. Truchado, E.C. Ferrando, P. Simon, A. Allende, G. Sanchez, SARS-CoV-2 RNA titers in wastewater anticipated COVID-19 occurrence in a low prevalence area, MedRxiv. (2020) 2020.04.22.20075200. https://doi.org/10.1101/2020.04.22.20075200.

[8] A. Nemudryi, A. Nemudraia, K. Surya, T. Wiegand, M. Buyukyoruk, R. Wilkinson, B. Wiedenheft, Temporal detection and phylogenetic assessment of SARS-CoV-2 in municipal wastewater, MedRxiv. (2020) 2020.04.15.20066746. https://doi.org/10.1101/2020.04.15.20066746.

[9] V.M. Corman, O. Landt, M. Kaiser, R. Molenkamp, A. Meijer, D.K.W. Chu, T. Bleicker, S. Brünink, J. Schneider, M.L. Schmidt, D.G.J.C. Mulders, B.L. Haagmans, B. van der Veer, S. van den Brink, L. Wijsman, G. Goderski, J.-L. Romette, J. Ellis, M. Zambon, M. Peiris, H. Goossens, C. Reusken, M.P.G. Koopmans, C. Drosten, Detection of 2019 novel coronavirus (2019-nCoV) by real-time RT-PCR, Eurosurveillance. 25 (2020). https://doi.org/10.2807/1560-7917.ES.2020.25.3.2000045.

[10] R.H. Kutner, X.-Y. Zhang, J. Reiser, Production, concentration and titration of pseudotyped HIV-1-based lentiviral vectors, Nat. Protoc. 4 (2009) 495–505. https://doi.org/10.1038/nprot.2009.22.

